# Hippocampal Glutamate, Resting Perfusion and the Effects of Cannabidiol in Psychosis Risk

**DOI:** 10.1101/2023.03.02.23286709

**Authors:** Cathy Davies, Matthijs G. Bossong, Daniel Martins, Robin Wilson, Elizabeth Appiah-Kusi, Grace Blest-Hopley, Paul Allen, Fernando Zelaya, David J. Lythgoe, Michael Brammer, Jesus Perez, Philip McGuire, Sagnik Bhattacharyya

**Affiliations:** Department of Psychosis Studies, Institute of Psychiatry, Psychology & Neuroscience, King’s College London, London, UK; Department of Neuroimaging, Institute of Psychiatry, Psychology & Neuroscience, King’s College London, London, UK; Department of Psychiatry, University Medical Center Utrecht Brain Center, Utrecht University, Utrecht, The Netherlands; National Institute for Health Research (NIHR) Maudsley Biomedical Research Centre (BRC), South London and Maudsley NHS Foundation Trust, London, UK; School of Psychology, University of Roehampton, London, UK; Combined Universities Brain Imaging Centre, Royal Holloway University of London, UK; CAMEO Early Intervention Service, Cambridgeshire and Peterborough NHS Foundation Trust, Cambridge, UK; Institute of Biomedical Research (IBSAL), Department of Medicine, Universidad de Salamanca, Salamanca, Spain; Department of Psychiatry, University of Oxford, Oxford, UK; NIHR Oxford Health Biomedical Research Centre, Oxford, UK; Oxford Health NHS Foundation Trust, Oxford, UK

**Keywords:** magnetic resonance spectroscopy, clinical high risk for psychosis, at-risk mental state, cerebral blood flow

## Abstract

**Background:** Preclinical and human data suggest that the onset of psychosis involves hippocampal glutamatergic dysfunction, driving hyperactivity/hyperperfusion in a hippocampal-midbrain-striatal circuit. Whether glutamatergic dysfunction is related to cerebral perfusion in patients at Clinical High Risk (CHR) for psychosis, and whether cannabidiol (CBD) has ameliorative effects on glutamate or its relationship with blood flow remains unknown.

**Methods:** Using a double-blind, parallel-group design, 33 CHR patients were randomised to 600mg CBD or placebo; 19 healthy controls did not receive any drug. Proton magnetic resonance spectroscopy was used to measure glutamate concentrations in left hippocampus. We examined differences relating to CHR status (controls vs placebo), effects of CBD (placebo vs CBD) and linear between-group effects, such that placebo>CBD>controls or controls>CBD>placebo. We also examined group x glutamate x cerebral perfusion (measured using arterial spin labelling) interactions.

**Results:** Compared to controls, CHR-placebo patients had significantly lower hippocampal glutamate (p=.015) and a significant linear relationship was observed across groups, such that glutamate was highest in controls, lowest in CHR-placebo and intermediate in patients under CBD (p=.031). There was also a significant interaction between group (controls vs CHR-placebo), hippocampal glutamate and perfusion in the putamen and insula (p_FWE_=.012), driven by a strong positive correlation in the CHR-placebo group vs a negative correlation in controls.

**Conclusions:** Our findings suggest that hippocampal glutamate is lower in CHR patients and may be partially normalised by CBD treatment. Furthermore, we provide the first *in vivo* evidence of an abnormal relationship between hippocampal glutamate and resting perfusion in the striatum and insula in these patients.

## INTRODUCTION

Preclinical and human data suggest that the onset of psychosis involves hippocampal glutamatergic dysfunction, driving hyperactivity and hyperperfusion in a hippocampal-midbrain-striatal circuit.^1–3^ Specifically, preclinical models propose that hippocampal hyperactivity—from glutamatergic pyramidal cell disinhibition—leads to excess excitatory drive in projections to the midbrain-striatum, causing hyper-responsivity of midbrain dopamine neurons, striatal hyperdopaminergia and the emergence of psychotic-like phenotypes.^1,3–5^ Consistent with this, evidence from human studies suggests that across the psychosis continuum, patients have altered concentrations of hippocampal glutamate or Glx^6^ (a composite of glutamate and glutamine), hippocampal and striatal hyperperfusion^3,7,8^ and elevated striatal dopamine synthesis capacity.^9,10^ Importantly, these pathophysiological features appear to emerge prior to the onset of psychosis in people at Clinical High Risk (CHR), progressively worsening and/or spreading as they transition to full-blown psychosis.^2,3,8,10^

In CHR individuals, previous research has identified abnormal concentrations of hippocampal glutamate and/or Glx, although both increases (in genetic risk individuals^11^), decreases^12,13^ and sometimes no differences^14–17^ have been reported. Within CHR groups, hippocampal glutamate is greater in those with poor vs good outcomes^18^ and (Glx) may be related to symptom severity.^19^ Separately, elevated perfusion (or cerebral blood flow; CBF) has been observed in the hippocampus,^7,20^ striatum^7,21^ and prefrontal cortex^7^ in CHR patients, and there is some evidence that this may be associated with altered neurochemistry. For example, previous work has linked prefrontal GABA levels^22^ and striatal dopamine function^23^ to hippocampal perfusion in CHR patients, and anterior cingulate glutamate/Glx to hippocampal perfusion in people with high schizotypy.^24^ Although differences in hippocampal glutamate and regional CBF have been identified (separately) in prior CHR studies, whether and how these parameters are associated with each other, and whether any such relationship is abnormal in CHR patients, has yet to be directly tested. A deeper understanding of how hippocampal glutamatergic dysfunction is related to other pathophysiological features—particularly within the hippocampal-midbrain-striatal circuit—would not only enhance understanding of the mechanisms underlying psychosis risk, but may also illuminate novel targets for preventative treatments. Given the current lack of effective pharmacotherapies for CHR patients,^25,26^ this remains a critical research priority.

One of the most promising candidate treatments is cannabidiol (CBD), a phytocannabinoid constituent of the cannabis plant.^27^ In contrast to the psychotomimetic and potential anxiogenic effects^28–32^ of delta-9-tetrahydrocannabinol, the main intoxicating cannabinoid in cannabis, CBD is non-intoxicating and has anxiolytic^33,34^ and antipsychotic properties.^35–37^ CBD modulates brain activation in response to cognitive and emotional fMRI tasks, particularly in medial temporal cortex and striatal regions, in both healthy and established psychosis cohorts.^38–43^ In CHR patients, we previously demonstrated that a 600mg dose of CBD partially normalises hippocampal resting perfusion,^44[preprint]^ and mediotemporal and striatal function during various fMRI tasks,^45,46^ such that perfusion/activation in the CBD group was intermediate between that of healthy controls and CHR patients under placebo. Accumulating evidence further suggests that CBD may have effects on glutamate. In people with first-episode psychosis, we previously found that CBD increased hippocampal glutamate, an effect linked to the greater reduction of positive symptoms observed under its influence.^47^ Independent work shows that CBD modulates Glx in basal ganglia and prefrontal cortex across ASD and neurotypical individuals.^48^ Altogether, these findings suggest that CBD may have effects on glutamate and blood flow in humans, two pathophysiological features strongly implicated in psychosis onset. However, whether CBD can normalise glutamatergic dysfunction (or its relationship with blood flow) in CHR patients is yet to be examined.

To fill this gap in knowledge, we examined hippocampal glutamatergic dysfunction in the CHR state and the effects of CBD using Proton Magnetic Resonance Spectroscopy (^1^H-MRS) and three parallel groups: CHR patients randomised to 600mg CBD or placebo and healthy controls. We first established whether hippocampal glutamate levels are altered in CHR placebo patients relative to controls. We then tested our primary hypothesis that CBD would at least partially normalise alterations in glutamate levels, such that a significant linear relationship (placebo>CBD>controls, or controls>CBD>placebo) would exist across groups. Finally, to probe the broader mechanistic relevance of glutamatergic dysfunction, we examined whether the relationship between glutamate and regional CBF (measured using wholebrain Arterial Spin Labelling; ASL) differed between groups and assessed the effects of CBD on this interaction.

### PATIENTS & METHODS

#### Participants

The study received Research Ethics (Camberwell St Giles) approval and all participants provided written informed consent. Thirty-three antipsychotic-naive CHR^49^ individuals, aged 18–35, were recruited from early detection services in the United Kingdom (see Supplementary Methods). Nineteen age (within 3 years), sex and ethnicity-matched healthy controls were recruited locally. Exclusion criteria included history of psychotic or manic episode, current DSM-IV diagnosis of substance dependence, IQ<70, neurological disorder, and contraindication to magnetic resonance imaging or treatment with CBD. Participants were required to abstain from cannabis for 96h, other recreational substances for 2 weeks, alcohol for 24h and caffeine and nicotine for 6h before attending. A urine sample prior to scanning was used to screen for illicit drug use.

#### Design, Materials, Procedure

Using a randomised, double-blind, placebo-controlled, three-arm parallel-group design, CHR participants were randomised to a single oral 600mg dose of CBD (THC-Pharm, Germany) or a matched placebo capsule. Psychopathology was measured at baseline (before drug administration) using the Comprehensive Assessment of At-Risk Mental States (CAARMS)^49^ and State-Trait Anxiety Inventory-State Subscale. Following a standard light breakfast, participants were administered the capsule of CBD or placebo (at ∼11AM) and 180min later, underwent a battery of MRI sequences. Control participants were investigated under identical conditions but did not receive any drug. Plasma CBD levels were sampled at baseline and at 120 and 300min after drug administration.

#### Magnetic Resonance Imaging

All scans were conducted on a General Electric Signa HDx 3T MR system with an 8-channel head coil. A whole-brain 3D sagittal T1-weighted scan (TE=2.85ms; TR=6.98ms; TI=400ms; flip angle=11°, voxel size=1.0×1.0×1.2mm) was acquired for voxel planning, co-registration and spatial normalisation of the ASL data, and calculation of ^1^H-MRS voxel tissue content. ^1^H-MRS spectra were acquired in the left hippocampus (Figure 1) using conventional Point-Resolved Spectroscopy acquisition (PRESS; TR=3000ms; TE=30ms; 96 averages) in a 6-minute scan. We employed the standard GE PROBE (Proton Brain Examination) sequence, which uses a standardised, chemically selective suppression (CHESS) water suppression routine. Unsuppressed water reference spectra (16 averages) were also acquired as part of the standard acquisition for subsequent eddy current correction and water scaling. Shimming was optimised, with auto-prescan performed twice before each scan. Using standardised protocols, the hippocampal voxel (right-left, anterior-posterior, superior-inferior: 20×20×15mm) was prescribed from the structural T1-weighted scan. Structural T1-weighted images were segmented using Statistical Parametric Mapping (SPM8) to enable calculation and correction for ^1^H-MRS voxel tissue content (Supplementary Methods). CBF was measured using 3D pseudo-Continuous ASL with acquisition parameters and preprocessing procedures in line with previous studies, as detailed in the Supplementary Methods.

**FIGURE 1.**
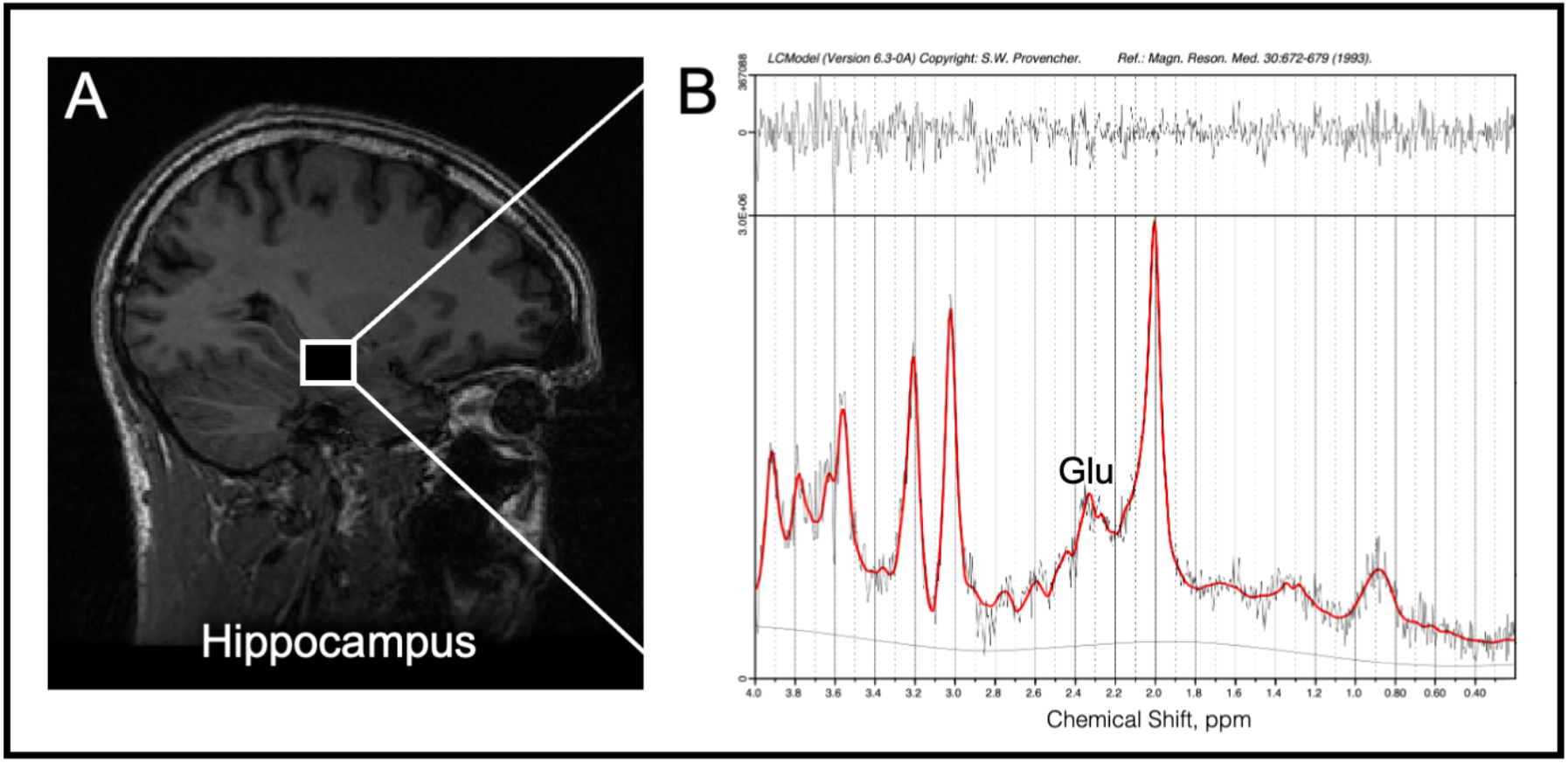
Example ^1^H-MRS voxel positioning and spectra in left hippocampus. In panel **A**, example voxel placement in the left hippocampus is indicated by the box. In panel **B**, ^1^H-MRS spectrum obtained (black line) from the voxel in A and the overlay of the spectral fit (red line). Glu indicates glutamate and ppm, parts per million.

#### ^1^H-MRS Data Processing

Spectra were analysed using LCModel/6.3-0A^50^ using the standard basis set of 16 metabolites (Supplementary Methods). Poorly fitted metabolite peaks (Cramer-Rao minimum variance bounds [CRLB] >20% as reported by LCModel) were excluded from further analysis. Water-scaled glutamate (primary outcome), glutamate plus glutamine (Glx), myo-inositol, creatine, choline, and N-acetylaspartate values were then corrected for voxel tissue composition (see Supplementary Methods). Spectral quality was further assessed using signal-to-noise ratio (SNR) and spectral linewidths (full width at half-maximum; FWHM).

### Statistical Analysis

Statistical analyses of ^1^H-MRS and other non-imaging data were performed in SPSS/27. Pairwise differences in clinical and demographic variables were examined using independent t-tests for continuous data and chi-square tests for categorical data. Potential group differences in data quality, including FWHM, SNR, CRLB and voxel tissue proportions were examined using independent t-tests (pairwise), and one-way ANOVA for any differences between groups. Before testing our primary hypothesis, we used independent samples t-tests to first establish (a) whether hippocampal glutamate levels were altered in the CHR placebo group vs controls, and (b) whether CHR patients under a single dose of CBD had altered hippocampal glutamate levels compared to CHR patients under placebo. Then, to test our primary hypothesis that glutamate levels in the CBD-treated group would be intermediate between that of the healthy control and placebo-treated group, we examined whether a linear relationship (controls>CBD>placebo; or placebo>CBD>controls) existed across groups, using one-way ANOVA and unweighted polynomial contrasts for linear degree. In the case of unequal variances, the same ANOVA was run using manual group weights (coefficients -1, 0, 1) which provides statistics (t-distribution) without assuming equal variances. Exploratory analyses of the secondary metabolites (Glx, myo-inositol, creatine, choline and N-acetylaspartate) were conducted using the same tests (described above) as for glutamate. The influence of individual metabolite values whose corresponding z-scores were >3 or <-3 (which may indicate outliers) were investigated by re-running statistical tests without them in sensitivity analyses. Significance was set at p<.05 (two-tailed).

#### Glutamate x CBF x Group Interactions

Group differences in the relationship between hippocampal glutamate and cerebral blood flow (glutamate x CBF x group interactions) were analysed using SPM12 in Matlab/R2018b. Using CBF as the dependent variable, glutamate levels were entered as a covariate of interest in independent t-tests for the two pairwise contrasts (placebo vs control; CBD vs placebo), and a flexible factorial ANCOVA model for the linear between-group analyses. In line with previous CHR studies of CBF,^7,20^ mean-centred age, gender, smoking status and years of education (the latter included due to significant group differences in our sample), as well as mean global CBF, were entered as nuisance covariates. We conducted a wholebrain search using an explicit grey matter mask (MNI152, thresholded at >.50) and cluster-level inference (cluster-forming threshold p<.005; clusters reported as significant at p<.05 using FWE cluster correction in SPM).

## RESULTS

There were no between-group differences in the majority of demographic and baseline clinical characteristics, except for fewer years of education in the placebo group relative to controls (Table 1), as reported in our previous publications.^45,46,51^ In the CBD group, mean plasma CBD levels were 126.4nM (SD=221.8) and 823.0nM (SD=881.5) at 120 and 300 min after drug intake, respectively. ^1^H-MRS data were available for all participants. ASL data were available for n=14 in the placebo group, n=14 in the CBD group and n=19 healthy controls.

**TABLE 1.** Sociodemographic and Clinical Characteristics at Baseline

| Characteristic |  | CBD<br>(n = 16) | Placebo<br>(n = 17) | Control<br>(n = 19) <sup>1</sup> | Pairwise Comparison |  |
| --- | --- | --- | --- | --- | --- | --- |
|  |  |  |  |  | Control vs<br>Placebo | Placebo<br>vs CBD |
| Age, years; mean (SD) |  | 22.7 (5.08) | 24.1 (4.48) | 23.9 (4.15) | p=.91 <sup>2</sup> | p=.42 <sup>2</sup> |
| Sex, N (%) male |  | 10 (62.5) | 7 (41.2) | 11 (57.9) | p=.32 <sup>3</sup> | p=.22 <sup>3</sup> |
| Ethnicity,<br>N (%) | White | 10 (62.5) | 7 (41.2) | 11 (57.9) | p=.59 <sup>3</sup> | p=.43 <sup>3</sup> |
|  | Black | 2 (12.5) | 5 (29.4) | 5 (26.3) |  |  |
|  | Asian | 0 (0) | 1 (5.9) | 0 (0) |  |  |
|  | Mixed | 4 (25) | 4 (23.5) | 3 (15.8) |  |  |
| Education, years; mean (SD) |  | 14.4 (2.71) | 12.6 (2.76) | 16.9 (1.58) | p<.001 <sup>2</sup> | p=.06 <sup>2</sup> |
| Handedness, N (%) right |  | 14 (87.5) | 17 (100) | 18 (94.7) | p=.37 <sup>3</sup> | p=.16 <sup>3</sup> |
| CAARMS Positive, mean (SD) |  | 40.19 (20.80) | 42.94 (29.47) | NA | NA | p=.76 <sup>2</sup> |
| CAARMS Negative, mean (SD) |  | 23.25 (16.49) | 28.41 (20.49) | NA | NA | p=.43 <sup>2</sup> |
| STAI-S, mean (SD) |  | 40.31 (9.07) | 38.94 (10.18) | NA | NA | p=.69 <sup>2</sup> |
| Urine drug<br>screen<br>results,<br>N (%) | Clean | 10 (63) | 8 (47) | NA <sup>4</sup> | Not<br>compared <sup>1</sup> | p=.45 <sup>3</sup> |
|  | THC | 2 (13) | 5 (29) | NA <sup>4</sup> |  |  |
|  | Morphine | 1 (6) | 0 (0) | NA <sup>4</sup> |  |  |
|  | Benzodiazepines | 0 (0) | 1 (6) | NA <sup>4</sup> |  |  |
|  | PCP | 0 (0) | 1 (6) | NA <sup>4</sup> |  |  |
|  | Missing | 3 (19) | 2 (12) | NA <sup>4</sup> |  |  |
| Current nicotine use, N (%) yes |  | 9 (56) | 5 (29) | 2 (11) | p=.15 <sup>3</sup> | p=.12 <sup>3</sup> |
| Current alcohol use, N (%) yes |  | 11 (69) | 10 (59) | NA | Not<br>compared <sup>1</sup> | p=.59 <sup>3</sup> |
| Lifetime cannabis use, N (%)<br>yes |  | 15 (94) | 17 (100) | NA <sup>5</sup> | Not<br>compared <sup>1</sup> | p=.48 <sup>3</sup> |
| Current cannabis use, N (%) yes |  | 7 (44) | 7 (41) | NA <sup>5</sup> | Not<br>compared <sup>1</sup> | p=.88 <sup>3</sup> |
| Cannabis<br>use<br>frequency,<br>N (%) | More than once a<br>week | 11 (69) | 12 (71) | NA | Not<br>compared <sup>1</sup> | p=.38 <sup>3</sup> |
|  | Once/twice<br>monthly | 1 (6) | 3 (18) | NA |  |  |
|  | Few times a year | 2 (13) | 0 (0) | NA |  |  |
|  | Only once/twice<br>lifetime | 1 (6) | 2 (12) | NA |  |  |

**Abbreviations:** CAARMS, Comprehensive Assessment of At-Risk Mental States; CBD, cannabidiol; CHR, Clinical High Risk for Psychosis; N, number of subjects; NA, not applicable; PCP, phencyclidine; STAI-S, State-Trait Anxiety Inventory-State Subscale; THC, Δ9-tetrahydrocannabinol. <sup>1</sup>Controls were selected to have minimal drug use and hence were not compared with CHR participants on these parameters; <sup>2</sup>Independent t-test; <sup>3</sup>Pearson chi-squared test; <sup>4</sup>Controls tested negative on urine drug screen for all substances tested; <sup>5</sup>Cannabis use less than 10 times lifetime (no current users).

### ^1^H-MRS data quality

Representative spectra for the hippocampal voxel is provided in Figure 1. Spectra were of good quality: aside from omission of choline data for two CHR subjects (see Table 2) due to CRLB>20%, no glutamate or other metabolite data were excluded. No significant differences in spectral quality nor voxel tissue content were observed between groups (Table 2). All metabolite values fell within a z-score of +/- 3.

**TABLE 2.** Spectral and structural voxel data. Mean ± SD estimates of linewidths, signal-to-noise ratios, CRLB, and voxel proportions of white matter, grey matter and CSF in the hippocampus across the three groups.

|  | Control<br>(n = 19) | Placebo<br>(n = 17) | CBD<br>(n = 16) | Control vs<br>Placebo | Placebo<br>vs CBD | ANOVA |
| --- | --- | --- | --- | --- | --- | --- |
| **Spectral and structural voxel data** |  |  |  |  |  |  |
| FWHM | 0.07 ± 0.02 | 0.07 ± 0.01 | 0.07 ± 0.01 | p=.84 | p=.18 | p=.50 |
| SNR | 12.53 ± 2.57 | 13.53 ± 2.40 | 13.38 ± 2.13 | p=.24 | p=.85 | p=.40 |
| WM | 0.33 ± 0.07 | 0.33 ± 0.06 | 0.36 ± 0.07 | p=.94 | p=.16 | p=.27 |
| GM | 0.63 ± 0.06 | 0.63 ± 0.05 | 0.61 ± 0.07 | p=.96 | p=.30 | p=.48 |
| CSF | 0.04 ± 0.02 | 0.04 ± 0.01 | 0.03 ± 0.02 | p=.63 | p=.09 | p=.07 |
| **Cramér Rao Lower Bounds (CRLB)** |  |  |  |  |  |  |
| Glu | 9.53 ± 2.06 | 9.76 ± 1.68 | 9.81 ± 2.61 | p=.71 | p=.95 | p=.91 <sup>b</sup> |
| Glx | 10.26 ± 2.10 | 10.29 ± 2.14 | 10.38 ± 3.30 | p=.97 | p=.93 | p=.99 <sup>b</sup> |
| NAA | 4.21 ± 1.32 | 3.88 ± 0.70 | 3.50 ± 0.63 | p=.35 | p=.11 | p=.09 <sup>b</sup> |
| Cho<sup>a</sup> | 4.00 ± 1.00 | 3.94 ± 0.77 | 3.93 ± 0.80 | p=.84 | p=.99 | p=.97 |
| ml | 5.58 ± 1.22 | 5.76 ± 1.20 | 5.94 ± 2.11 | p=.65 | p=.77 | p=.79 |
| Cre | 3.89 ± 0.66 | 4.06 ± 0.66 | 3.88 ± 0.72 | p=.46 | p=.45 | p=.69 |

*Abbreviations:* FWHM, full width at half-maximum (linewidth) in ppm (parts per million); WM, white matter; GM, grey matter; CSF, cerebrospinal fluid; CRLB, Cramér Rao Lower Bounds; SNR, signal-to-noise ratio; Glu, Glutamate; Glx, Glutamate + Glutamine; NAA, N-acetylaspartate; Cho, Choline; ml, myo-Inositol; Cre, Creatine. <sup>a</sup>Choline data for two CHR subjects (one from each of the CBD and placebo groups) was omitted due to CRLB >20%.
<sup>b</sup>Welch's test due to inhomogeneity of variances

### Hippocampal Glutamate – Pairwise effects of CHR status and CBD

Compared to healthy controls, CHR patients in the placebo group had significantly lower hippocampal glutamate (mean±SD in controls= 8.41 ± 1.27; placebo= 7.42 ± 1.02; t(34)= 2.55, p=.015) (Fig 2A, Table 3). Although hippocampal glutamate levels were numerically higher in the CBD (7.83 ± 1.67) vs placebo (7.42 ± 1.02) group, the pairwise difference was not significant (p=.41; Table 3).

**FIGURE 2.**
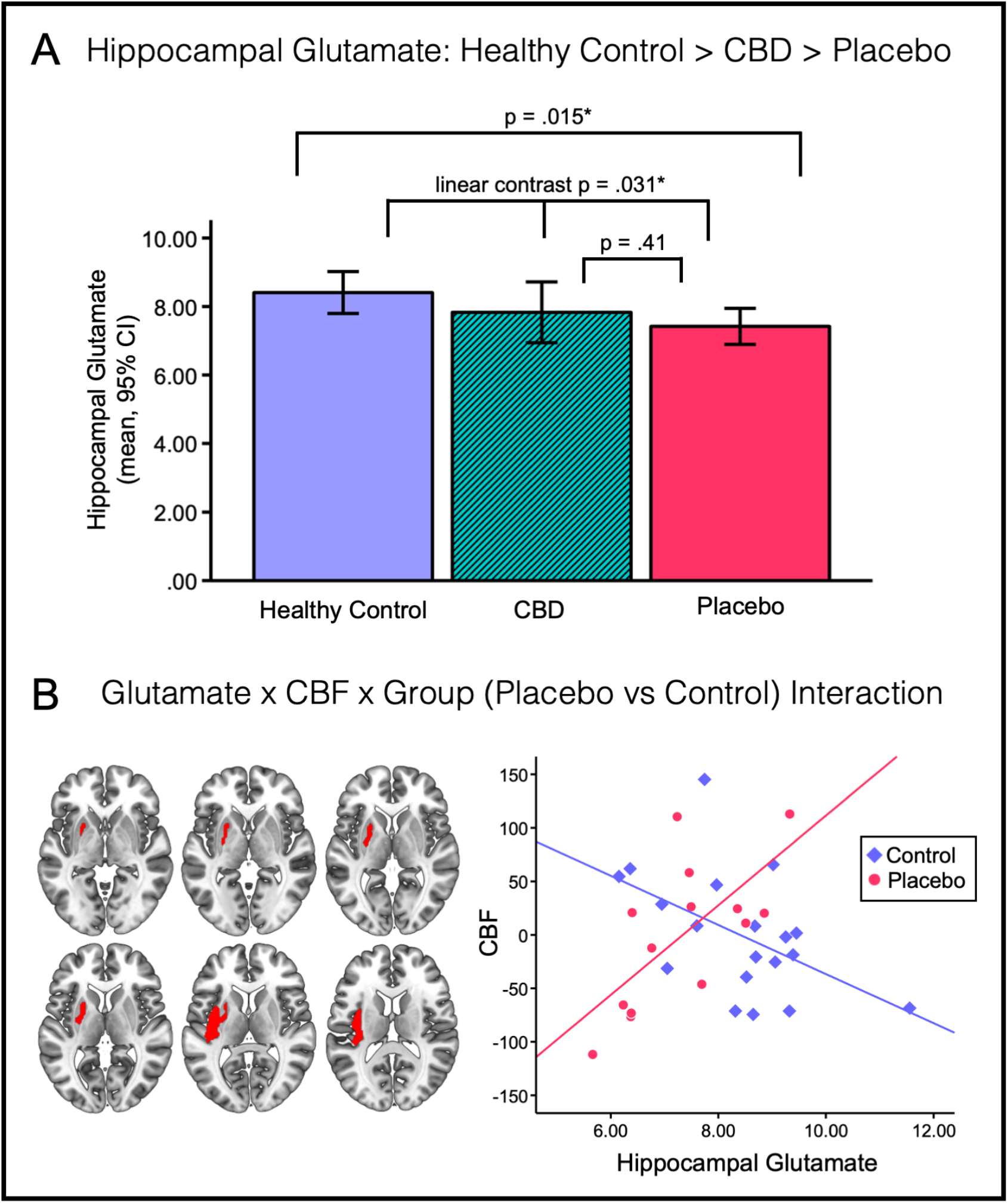
**(A)** Mean hippocampal glutamate levels across the three groups. Healthy controls had significantly higher glutamate relative to CHR-placebo patients (p=.015) and there was a significant linear relationship across groups (such that controls>CBD>placebo; p=.031). In the left panel of (**B**), axial sections showing the significant cluster (in red) identified in the glutamate x CBF x group (placebo vs control) interaction analyses (peak MNI X/Y/Z=-38/-12/10, T(24)=5.63, k=679, p_FWE_=.012). The right side of the brain is shown on the right of the images. In the right panel of (B), scatterplot depicting the relationship between CBF (in the putamen-insula cluster shown in the left panel) and hippocampal glutamate by group. This post-hoc analysis was used to determine the direction of the significant interaction: covariate-adjusted CBF values (mean CBF over all voxels, in arbitrary units) were extracted from the significant cluster (left panel) for each subject from the T-contrast using the MarsBaR toolbox in SPM.

**TABLE 3.**
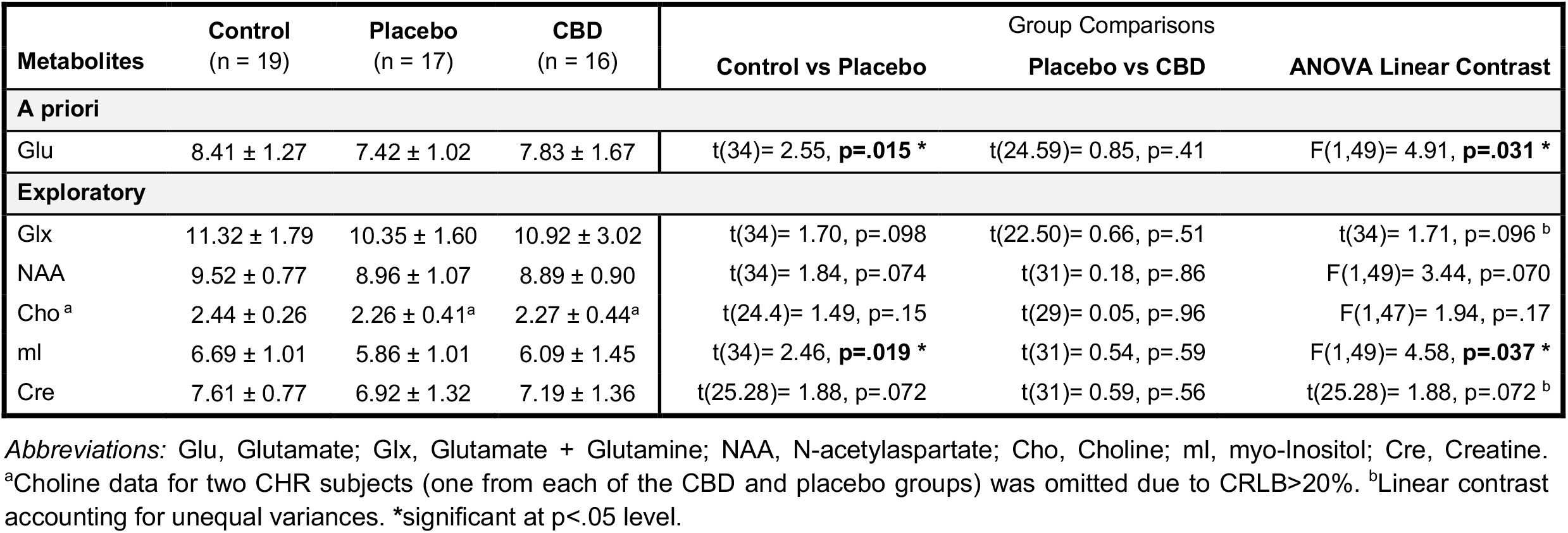
Tissue-corrected metabolite values in the hippocampus (mean ± SD) across the three groups, with pairwise comparisons (healthy control vs placebo; placebo vs CBD) and three-way contrasts for a between-group linear relationship (controls > CBD > placebo, or placebo > CBD > controls).

### Hippocampal Glutamate – Between-Group Linear Analyses

In our primary *a priori* analyses, we found a significant linear relationship across groups, such that hippocampal glutamate was highest in healthy controls, lowest in placebo-treated patients and intermediate in patients treated with CBD (ANOVA unweighted linear term F(1,49)= 4.91, p=.031) (Fig 2A, Table 3).

### Glutamate x CBF x Group Interactions

There was a significant interaction between group (control vs placebo), hippocampal glutamate and CBF in a cluster spanning the left putamen and insula (peak MNI coordinates X= -38 Y= -12 Z= 10, T(24)= 5.63, k=679, p_FWE_=.012). Post-hoc analysis demonstrated that this was characterised by a strong positive correlation in the placebo group (r=.68, p=.008, n=14) vs a negative correlation in healthy controls (r= -.51, p=.027, n=19) (Fig 2B). There were no significant interactions in the CBD vs placebo contrast nor the three-group linear analyses.

### Exploratory Effects on Other Metabolite Levels

Analysis of the secondary/exploratory metabolites revealed significantly lower myo-inositol in the placebo (M±SD= 5.86±1.01) relative to the control (6.69±1.01) group (t(34)= 2.46, p=.019). Although the direct pairwise CBD (6.09±1.45) vs placebo comparison for myo-inositol was not significant (p=.59), there was a significant linear relationship across groups, such that it was highest in healthy controls, lowest in placebo patients and intermediate in patients treated with CBD (ANOVA unweighted linear term F(1,49)= 4.58, p=.037). There were no other significant effects in pairwise or linear contrast analyses for any of the other metabolites (Glx, N-acetylaspartate, choline, creatine)(Table 3). Note that these results were not corrected for multiple testing as they are exploratory outcomes only.

## DISCUSSION

This is the first study to investigate the effects of CBD on hippocampal neurochemistry—and its association with regional cerebral perfusion—in people at CHR for psychosis. We first established that hippocampal glutamate levels are lower in CHR patients under placebo relative to healthy controls. To examine our primary hypothesis that CBD would at least partially normalise any such glutamatergic alterations, we then tested for a linear relationship across groups. In line with our predictions, our first major finding was that hippocampal glutamate levels in the CBD-treated CHR group were significantly intermediate between those observed in the placebo group and healthy controls. Finally, we provide the first *in vivo* evidence that hippocampal glutamate levels are abnormally associated with perfusion in the striatum and insula in CHR patients relative to controls. Together, these results provide novel insights on the neurobiological mechanisms underlying psychosis risk and suggest that CBD may partially normalise glutamatergic dysfunction in these patients.

### Hippocampal Glutamate is Lower in CHR Patients vs Controls

Our finding that hippocampal glutamate was lower in the CHR placebo group vs controls is consistent with several previous studies demonstrating lower hippocampal Glx^13^ and glutamate^12^ (sometimes only at trend-level)^13,52^ in these patients, as well as negative associations between hippocampal glutamate, striatal dopamine and symptom severity, particularly in those who go on to transition^53^ (although see^14^). However, other studies have found increased hippocampal Glx (albeit in those with genetic risk),^11^ no differences,^14–17^ or differences only *within* CHR patients based on poor vs good clinical outcomes.^18^ The reasons for the disparity in the presence and/or direction of results are unclear, but sample characteristics^54^ or methodological factors such as voxel location, metabolite correction (for voxel tissue content vs creatine-scaled), as well as the heterogeneity inherent within CHR populations^55,56^ may contribute. These findings are compatible with the several meta-analyses that have synthesised this literature,^6,57,58^ which report numerically (but non-significantly) lower hippocampal glutamate levels in CHR individuals relative to controls (SMD[g]=-0.26, 95%CI: -0.56-0.04, p=.09).^57^ Nevertheless, it is also plausible that excess vs attenuated hippocampal glutamate exists in subsets of CHR individuals. For example, the largest study of hippocampal metabolites in CHR patients to date reported significantly lower hippocampal glutamate in those with good vs poor outcomes, and the point estimate in the good outcome subgroup was even numerically (albeit non-significantly) lower than that in healthy controls.^18^ Therefore, we cannot exclude that heterogeneity in CHR samples may account for at least some of the conflicting findings across studies. Together, our findings add to previous literature showing that hippocampal glutamate is abnormal in people at CHR. Going forward, consortia-scale studies are needed to examine whether subgroups based on hippocampal glutamate exist in CHR cohorts and to characterise their respective clinical and neurophysiological features.

### CBD may Increase Hippocampal Glutamate in CHR Patients

Our first major finding was that CHR patients treated with a single dose of CBD show intermediate levels of hippocampal glutamate relative to controls and patients under placebo. Although our study was cross-sectional with parallel groups, these results suggest that a single dose of CBD may partially normalise the altered glutamate levels we observed in CHR patients. Supporting this view, in our previous within-subject study in people with first-episode psychosis, we showed that a 600mg dose of CBD significantly increased hippocampal glutamate relative to placebo.^47^ Moreover, CBD was associated with a significantly greater decrease in symptom severity, and a significant inverse relationship was found between hippocampal glutamate and the severity of psychotic symptoms post-treatment.^47^ This suggests that the antipsychotic effects of CBD in patients with psychosis^36,37^ may be related to the increase in hippocampal glutamate observed under its influence.^47^ Outside of the hippocampus, CBD has been shown to increase Glx in basal ganglia but reduce Glx in prefrontal cortex across ASD and neurotypical individuals.^48^ Preclinical studies demonstrate that CBD can increase prefrontal glutamate in rodent depression models,^59^ although attenuated glutamate release from hippocampal synaptosomes has been observed in cocaine-induced seizure models.^60^ CBD may also act on excitation-inhibition balance via the GABAergic system.^61^ In humans, CBD increases GABA in basal ganglia and prefrontal cortex in controls, but decreases GABA in these regions in ASD individuals.^48^ Overall, previous work points to effects of CBD on the glutamate system but the regions implicated and the direction of effects are somewhat mixed, potentially due to species-specific differences, the differential populations examined in humans and other methodological factors. Our results therefore extend the limited body of existing literature (so far conducted in people with established psychosis, ASD and neurotypical controls) by showing that CBD may also modulate hippocampal glutamate in people at risk of psychosis, and in a direction indicative of normalisation. However, it should be noted that we did not find significant differences in the CBD vs placebo pairwise analysis. This may be due to the relatively modest sample sizes (n=17/16 per CHR group) and thus limited power for detecting effects of smaller magnitude. This lack of differences is perhaps unsurprising, since it would be unlikely that a single dose of CBD would fully normalise glutamatergic dysfunction in CHR patients. Therefore, with this in mind we tested our hypothesis of a partial normalisation effect of CBD directly using the three-way linear analyses, as in our previous publications in this sample.^45,46,51^ Our findings lend support to the idea that CBD may hold value as a potential therapeutic avenue to be pursued in further clinical studies. Future work employing a within-subject design, and studies administering CBD to a larger group of CHR patients repeatedly over longer durations are needed to further characterise the direct effects of CBD on glutamate in these patients.

### Abnormal Relationship Between Glutamate & Perfusion in CHR Patients

In exploring the broader mechanistic relevance of glutamatergic dysfunction, our second major—and novel—finding was of a significant group (control vs placebo) x glutamate x CBF interaction in a putamen-insula cluster, driven by a strong positive association in the CHR placebo group and a negative association in controls. Dysfunctional relationships between hippocampal and putaminal physiology are of particular interest as the striatum is a key node in circuit-based models of psychosis.^1^ These propose that hippocampal hyperactivity leads to excess excitatory drive in projections to the midbrain-striatum, striatal hyperdopaminergia and the emergence of psychotic symptoms.^1,4,5^ If, as our results in healthy controls suggest, the normative relationship is such that greater hippocampal glutamate is associated with lower striatal CBF, the positive association we observed in CHR patients could reflect a disruption in inhibitory/homeostatic mechanisms within this hippocampal-midbrain-striatal circuit. Our findings are thus broadly consistent with the aforementioned preclinical models.

In addition, our finding that glutamate was atypically related to perfusion in a cluster localised to the putamen and insula is interesting in light of the putamen hyperperfusion documented in two previous CHR studies.^7,21^ Increased putamen CBF in CHR patients^21^ also correlates with positive symptom severity, and lower striatal CBF at follow-up has been associated with greater longitudinal decreases in positive symptoms.^7^ Greater perfusion in the putamen has also been identified as a potential marker of genetic susceptibility for schizophrenia spectrum disorders in a neuroimaging twin study.^62^ In terms of the insula, perfusion abnormalities have not been definitively reported here in the CHR state, but a recent meta-analysis found conjoint reductions in CBF and glucose metabolism (indexing aberrant neurovascular coupling) within frontoinsular cortex in schizophrenia.^63^ In our previous study in the same sample, we focused exclusively on perfusion and found (during exploratory wholebrain analyses) significantly increased CBF in placebo-treated patients vs controls. This large cluster extended into the left putamen (but not the insula) and partially overlaps with the cluster found here.^44[preprint]^ The present findings therefore extend our prior work to collectively suggest that (a) hippocampal glutamate is lower, (b) CBF in (clusters spanning) the striatum is greater, and (c) that the relationship between hippocampal glutamate and striatal-insular perfusion is abnormal in CHR patients relative to controls. While previous work has found CHR-associated dysfunction in the relationship between prefrontal GABA and hippocampal blood flow,^22^ and between hippocampal glutamatergic metabolites and (i) striatal dopamine,^53^ (ii) hippocampal activation (by clinical outcomes),^64^ and (iii) hippocampal-striatal connectivity,^64^ the current study is the first to demonstrate an aberrant relationship between *hippocampal glutamate* and *striatal blood flow* in these patients. Given that hippocampal glutamatergic dysfunction is thought to drive hyperperfusion in the hippocampal-midbrain-striatal circuit, our results provide new empirical evidence of a potential link between these two pathophysiological features in CHR patients, which have so far only been reported in isolation. Our findings therefore provide novel insights on potential mechanisms underlying psychosis risk from a complementary angle to previous literature, and provide a starting point for future research to unpack the nature of these alterations on a more granular level.

### Limitations

Several potential limitations warrant consideration. First, our use of a parallel-group design means that the effects of CBD ascribed from the linear between-group analyses should be interpreted with caution, as it is possible—although we think unlikely, based on commensurate results in our previous within-subject study^47^—that they emerged due to random between-group variation, rather than effects of CBD. Future studies employing a within-subject design in CHR patients would address this issue. Second, CBF data were missing for several CHR subjects, which may have impacted the statistical power of the combined glutamate-CBF analyses. More generally, inherent limitations of ^1^H-MRS include the fact that the voxel size is large and concentration estimates reflect both intracellular and extracellular glutamate, without an ability to distinguish between glutamate involved in neurotransmission vs metabolism.^65^ Scanning at higher field strengths and advanced techniques (such as GluCEST) are now becoming available and will enable future research to more reliably separate spectral components.^66,67^ Finally, our study was designed and powered to detect neurophysiological rather than symptomatic effects. As such, future studies combining ^1^H-MRS, CBF and symptom outcomes in larger CHR samples are needed to establish whether CBD’s effects on glutamate or CBF are related to concomitant symptomatic improvement.

## Conclusion

In summary, we found that hippocampal glutamate is lower in CHR patients and may be partially normalised by a single dose of CBD. Furthermore, we provide the first *in vivo* evidence of an abnormal relationship between hippocampal glutamate and resting perfusion in the striatum and insula in this clinical group. Together, these results provide novel insights on the neurobiological mechanisms underlying psychosis risk and suggest that CBD warrants further investigation as a candidate novel treatment.

## Data Availability

Data produced in the present work are not openly available.

## ACKNOWLEDGEMENTS

The authors wish to thank the study volunteers, and the radiographers at the Centre for Neuroimaging Sciences, King’s College London, who carried out the MRI scans.

## AUTHOR CONTRIBUTIONS

Substantial contributions to conception and design (SB, PMG, PA, MGB, DL, FZ, MB), acquisition of data (RW, EAK, GBH), analysis and/or interpretation of data (CD, SB, MGB, DM), drafting the article (CD, SB) or revising it critically for important intellectual content (all authors), study supervision (SB), final approval of the version to be published (all authors).

