## Supplementary Material for "Hippocampal Glutamate, Resting Perfusion and the Effects of Cannabidiol in Psychosis Risk"

**Supplementary Methods**

- Participants
- Design, Materials, Procedure
- $^{1}$H-MRS Data Processing
- Arterial Spin Labelling Acquisition & Processing

### SUPPLEMENTARY METHODS

#### *Participants*

Thirty-three antipsychotic-naive CHR individuals, aged 18–35, were recruited from specialist early detection services in the United Kingdom. CHR status was determined using the Comprehensive Assessment of At-Risk Mental States (CAARMS) criteria.[1] Briefly, subjects met one or more of the following subgroup criteria: (a) attenuated psychotic symptoms, (b) brief limited intermittent psychotic symptoms (BLIPS, psychotic episode lasting <1 week, remitting without treatment), or (c) either schizotypal personality disorder or first-degree relative with psychosis, all coupled with functional decline.[1]

#### *Design, Materials, Procedure*

The 600mg dose of CBD was selected based on previous findings that doses of 600-800 mg/day are effective in established psychosis[2] and anxiety.[3] The 180 min interval between drug administration and MRI acquisition was selected based on previous findings describing peak plasma concentrations at 180 min following oral administration.[4,5]

#### *$^1$H-MRS Data Processing*

Spectra were analysed using LCModel/6.3-0A[6] using the standard basis set of 16 metabolites (L-alanine, aspartate, creatine, phosphocreatine, GABA, glucose, glutamine, glutamate, glycerophosphocholine [choline], glycine, myo-inositol, L-lactate, N-acetylaspartate, N-acetylaspartylglutamate, phosphocholine, and taurine) acquired at the same field strength (3T), localisation sequence (PRESS), and echo time (30ms) as the $^1$H-MRS spectra in the current study. Model metabolites and concentrations used in the basis set are detailed in the LCModel manual (http://s-provencher.com/pub/LCModel/manual/manual.pdf).

We calculated and corrected for $^1$H-MRS voxel tissue content using SPM8 and in-house scripts to (a) segment the T1-weighted structural images into grey matter, white matter, and cerebrospinal fluid (CSF) using SPM8 in Matlab R2017a, (b) locate and map the coordinates of each voxel to the segmented T1 images, and (c) provide the tissue content proportions. Metabolite values were corrected for voxel tissue content using the formula: $M_{corr} = M \times ([GM \times 1.21] + WM + [CSF \times 1.55]) / (WM + GM)$, where M is the uncorrected metabolite value and GM/WM/CSF are proportions of grey matter, white matter and CSF, respectively. The formula assumes a CSF water concentration of 55,556 mol/m$^3$ and the LCModel default brain water concentration of 35,880 mol/m$^3$.[7,8] Apart from assuming $T_2$ = 80 ms for tissue water, no corrections were applied for metabolite and water relaxation times.

***Arterial Spin Labelling Acquisition & Processing***

*ASL Image Acquisition*

For ASL image registration, a high resolution T2-weighted Fast Spin Echo (FSE) image (TE= 54.58ms, TR= 4380ms, Flip angle= 90deg, FoV= 240, Matrix size= 320 x 320, slice thickness= 2mm, 72 spatial locations) was acquired and used alongside the T1-weighted Spoiled Gradient Recalled (SPGR) images (detailed in the main text). Resting Cerebral Blood Flow (CBF) was measured using 3D pseudo-Continuous Arterial Spin Labelling (CASL) scans acquired with a 3D Fast Spin Echo (FSE) spiral multi-shot readout, following a post-labelling delay of 1.5s. The spiral acquisition used a short (10ms) TE, and 8 spiral arms (interleaves) with 512 points in each arm. FSE TE= 32.26ms, TR = 5500ms. 64 slices of 3mm thickness were obtained and the in-plane FoV was 240×240mm. Three pairs of tagged-untagged images were collected. The whole ASL pulse sequence, including the acquisition of calibration images, was performed in 6:08min.

*ASL Image Processing*

Data were preprocessed using FMRIB Software Library (FSL) 6.0.2 using the following procedure: (1) T1 and T2 images were skull-stripped and corresponding brain-only binary masks created; (2) original CBF images were coregistered to the T2 images and (3) multiplied by the binary T2 mask to create a skull-stripped CBF image in T2 space; (4) skull-stripped T2 was coregistered to skull-stripped T1; (5) skull-stripped T1 was first linearly coregistered to the MNI152 T1 2mm brain template, before non-linear registration (FNIRT) of the original T1 to MNI space; (6) original T2 images were registered to the MNI template (via T1 space) in a single concatenated step, using the T2-to-T1 transformation matrix (from step 4) and T1-to-MNI warp (from step 5); (7) skull-stripped CBF images (already in T2 space) were registered to the MNI template using the concatenated procedure in step 6; (8) normalised CBF images were spatially smoothed with a 6mm Gaussian kernel. All images were visually inspected for preprocessing errors.
